# Covering all the BASEs? Barriers and facilitators to STI testing among international students living in England

**DOI:** 10.64898/2026.10.29.26366809

**Authors:** Cristina Santamaria-Plaza, Carina Hörst, Avelie Stuart, Prarthana Narayanan, Michelle J Cole, Katy Sinka, Catherine M. Lowndes, Hamish Mohammed

## Abstract

**Background:** The emergence in England of ceftriaxone-resistant *Neisseria gonorrhoeae*, the bacteria which causes gonorrhoea, is a public health concern. Most cases of ceftriaxone-resistant gonorrhoea in England are travel-associated. We assessed the barriers and facilitators to sexually transmitted infection testing among international students living in England.

**Methods:** We conducted semi-structured interviews (March-August 2025) with international students attending a university in London. We recruited students through various university networks and snowball sampling. We used the Capability, Opportunity and Motivation (COM-B) model and conducted a thematic analysis.

**Results:** We conducted 15 interviews. Most participants were from China (n=8) and Nigeria (n=2), with other participants from countries in Europe, Asia, the Middle East and the Americas. Most participants were 25-34 years old and in a postgraduate programme. Two-thirds of our sample had previously been tested for STIs (7 in England, and 3 in their home country).

STI language and knowledge were key psychological capability barriers to STI testing, awareness of STI testing options was a facilitator. Under physical capability, no STI symptoms influenced testing for some, but prompted testing for others. Belief in confidentiality, privacy, and anonymity of testing, alongside perceived social and individual benefits of STI testing, were identified as facilitators under reflective motivation. Perceived risk varied based on relationship status. Anticipated judgement and stigma were the main social opportunity barriers, while trust in health professionals was the main facilitator. Fragmented services and poor proximity were key physical opportunity barriers; perceived availability of services and access to free, straightforward testing were facilitators.

**Conclusions:** We integrated these findings into national STI public health messaging targeted at university students in the England. The findings can be used to create culturally appropriate resources to raise awareness of STI-related symptoms and, among those with a need, promote STI testing.

## Introduction

*Neisseria gonorrhoeae*, the sexually transmitted bacteria which causes gonorrhoea, is designated a high-priority pathogen by the World Health Organization due to widespread antimicrobial resistance (AMR). In England, the emergence of ceftriaxone-resistant *N. gonorrhoeae* is a public health concern as ceftriaxone is the last remaining antibiotic currently available for use as empirical monotherapy. As of August 2026, most of the 88 cases of ceftriaxone-resistant gonorrhoea detected in England have been travel-associated; mainly from individuals, including international students, with links to the Asia-Pacific, the region with the highest prevalence of ceftriaxone-resistant *N. gonorrhoeae* globally (1–3).

England is a top destination for international students, with China and Nigeria as the main countries of origin. Students aged 21-24 consistently represented the largest age group among international students enrolled in UK higher education between 2020/21 and 2024/25 accounting for 42% of enrolments in 2024/25 (4, 5). In England, sexual health services are provided free at the point of delivery to everyone irrespective of nationality or visa status – and this includes sexually transmitted infection (STI) testing at sexual health services (SHS), or through online postal self-sampling kits for STIs. While there have been some research studies with students examining barriers and facilitators to STI testing, they have predominantly been among domestic students (6–8); due to the unique set of cultural and structural challenges faced by international students, these findings may not be generalisable to international students. Though there is some existing evidence that indicates that knowledge, attitudes, social and structural factors, as well as inequalities in access to SHS can affect the uptake of STI testing among young people (6, 8–11), including international students (12–16), this area is under-researched in England.

Rapid detection and contact tracing are essential for the control of ceftriaxone-resistant *N. gonorrhoeae* strains, so it is important that international students, who may be unfamiliar with sexual healthcare delivery in England, access STI testing when needed. We assessed the barriers and facilitators to STI testing among international students living in England and developed recommendations to mitigate the barriers.

## Methods

### Sampling strategy

International students from a university in London with a high proportion of international students were recruited for this study. Participants were deemed eligible if they were current or prior students, aged between 18 and 34 years, had resided in England for at least 6 months up to a maximum of 5 years, and were fluent in English. Participants did not need to have had an STI test in England before, nor did they need to have had any sexual experiences with others before the interview. We aimed to recruit 20 participants, as this is a sample size that is used in similar research studies (8, 14, 17). Data were reviewed throughout data collection to assess whether saturation has been reached, defined as the point at which no new themes or substantive insights were identified (18). Data collection was halted after 15 interviews due to logistical reasons. However, all our participants shared a common background – being an international student in England – which was critical to our research question, and we are confident that our sample is therefore specific enough to gain answers to our research question (19).

### Design and theoretical framework

We performed one-on-one virtual semi-structured interviews, using a topic guide informed by the COM-B model (20). The COM-B model is widely used in public health research to explain behaviour and covers the domains of capability (“C”; physical and psychological), opportunity (“O”; social and physical), and motivation (“M”; automatic and reflective). It is suitable for identifying barriers and facilitators to a target behaviour. The framework is linked to intervention frameworks, such as the Behaviour Change Wheel (20).

### Data collection

We recruited participants through university contacts who digitally distributed the recruitment materials to their students via newsletters, mailing lists, and social media. We also used a snowball sampling approach whereby students recruited for the study identified other participants through their social networks. Interested participants completed an intake survey to assess eligibility. Eligible participants were provided with a consent form and a participant information sheet.

CS and CH conducted the interviews guided by the topic guide (S1 Topic guide in S1 Text). Audio interviews were conducted via Microsoft Teams and ranged in length from 30-60 minutes. Interviews were audio recorded and transcribed verbatim. Interviews took place between 18 March and 8 August 2025. Participants were compensated with a £40 voucher for an online multi-retailer website. Participant contact information was kept separately from recordings, transcripts, and interview notes, and names were deleted from transcripts. Interview recordings were temporarily stored on a secure UKHSA drive and deleted immediately from any local drives after upload.

### Data analysis

We conducted a thematic analysis using the COM-B model. We used NVivo 14 for data analysis, following a hybrid approach to coding (21) and a 6-step analysis:

1. CS familiarised herself with the interview data.
2. CS generated an initial codebook based on three transcripts. The codes were reviewed by CH. CS deductively coded the data to the COM-B domains as either barriers, facilitators, recommendations, other and/or general context. Data was assigned to two or more COM-B domains where relevant. In some instances, participants described factors that made the target behaviour more difficult or easier but did not discourage or facilitate it. These accounts were distinguished from those classified as barriers or facilitators to support a more precise mapping to the Behaviour Change Wheel. Mixed accounts, in which the same factor was described as facilitating and preventing STI testing by the same individual, were identified and accounted for during coding. Codes were inductively generated.
3. To ensure the reliability of the codes, the codebook was iteratively reviewed by CH and AS.
4. The final codebook was exported to Microsoft Excel, and codes were visually clustered into themes. Themes were cross-checked by CH, AS, PN, and HM, and were discussed among all authors. Some themes encompassed factors that both facilitated and discouraged STI testing and were therefore retained as mixed themes.
5. While all themes in qualitative analyses are considered meaningful irrespective of frequency, we applied pragmatic thresholds to identify higher-priority themes to inform intervention recommendations. Themes including accounts of eight or more participants were classified as high priority, and those of between five and eight participants as medium priority. The results section is comprised solely of these themes. The low-priority themes (under 5 participants) can be found in S1 Table.
6. A thematic map was generated to visualise the themes and condense them into larger narratives (or overarching themes).

### Patient and public involvement and engagement

Participant-facing materials were reviewed by two former international students, one each from China and Nigeria – representing the two highest proportions of nationalities among our sample. Feedback was reviewed and integrated into the final products. The interview guide was piloted with two UK Health Security Agency (UKHSA) employees with who have lived experience of being international students in the UK.

### Ethical considerations

This study was approved by the UKHSA Research Ethics and Governance Group (REGG) no. R&D 603. The research team also obtained permission from the ethics board of the target university in London (name omitted to protect participants’ confidentiality) to recruit among their international students. Written consent from all participants was obtained prior to conducting the interviews.

### Researcher characteristics and reflexivity

The study team comprised individuals with diverse disciplinary and methodological backgrounds including behavioural science, epidemiology, and qualitative research. The interdisciplinary team composition allowed for open discussion about the interpretation of the data through multidisciplinary lenses. None of the members were international students at the time of the study, therefore, the data was interpreted from an external perspective.

Despite not having lived experience as an international student in the UK, the lead author’s (CS) own experience of adjusting to a new culture and navigating unfamiliar health systems created potential points of connection with participants. This could have resulted in greater attention being given to experiences that resonated with her own. Additionally, CS’s position within a public health organisation could have influenced what participants felt comfortable sharing. Discussions with the wider team were used to challenge assumptions and consider alternative interpretations.

## Results

Table 1 describes the main demographic characteristics of participants. We recruited a total of 15 participants. Participants were mostly from China (eight participants) and Nigeria (two participants) with other participants from countries in Europe, Asia, the Middle East and the Americas (one each). Most participants were between the ages of 25 to 34 years and were enrolled in a post-graduate degree programme. One third of the participants were enrolled in a health-related degree. Two thirds of our sample had previously been tested for STIs, including in England.

**Table 1.** Participant demographics.

| **Age group (years)** |  |
|---|---|
| 18-24 | 4 |
| 25-34 | 11 |
| **Sex** |  |
| Female | 8 |
| Male | 7 |
| **Level of study** |  |
| Undergraduate | 1 |
| Postgraduate | 13 |
| Alumni | 1 |
| **Test status** |  |
| Tested in the UK | 7 |
| Tested in home country | 3 |
| Never tested | 5 |
| **Field of study** |  |
| Social Sciences | 8 |
| Health | 5 |
| Other | 2 |

Themes were grouped into 5 overarching narratives and are presented by COM-B domain (Fig 1). Quotes that clearly and strongly expressed a given theme were selected as representative quotes. Quotes in some cases could be coded to more than one domain.

**Fig 1.**
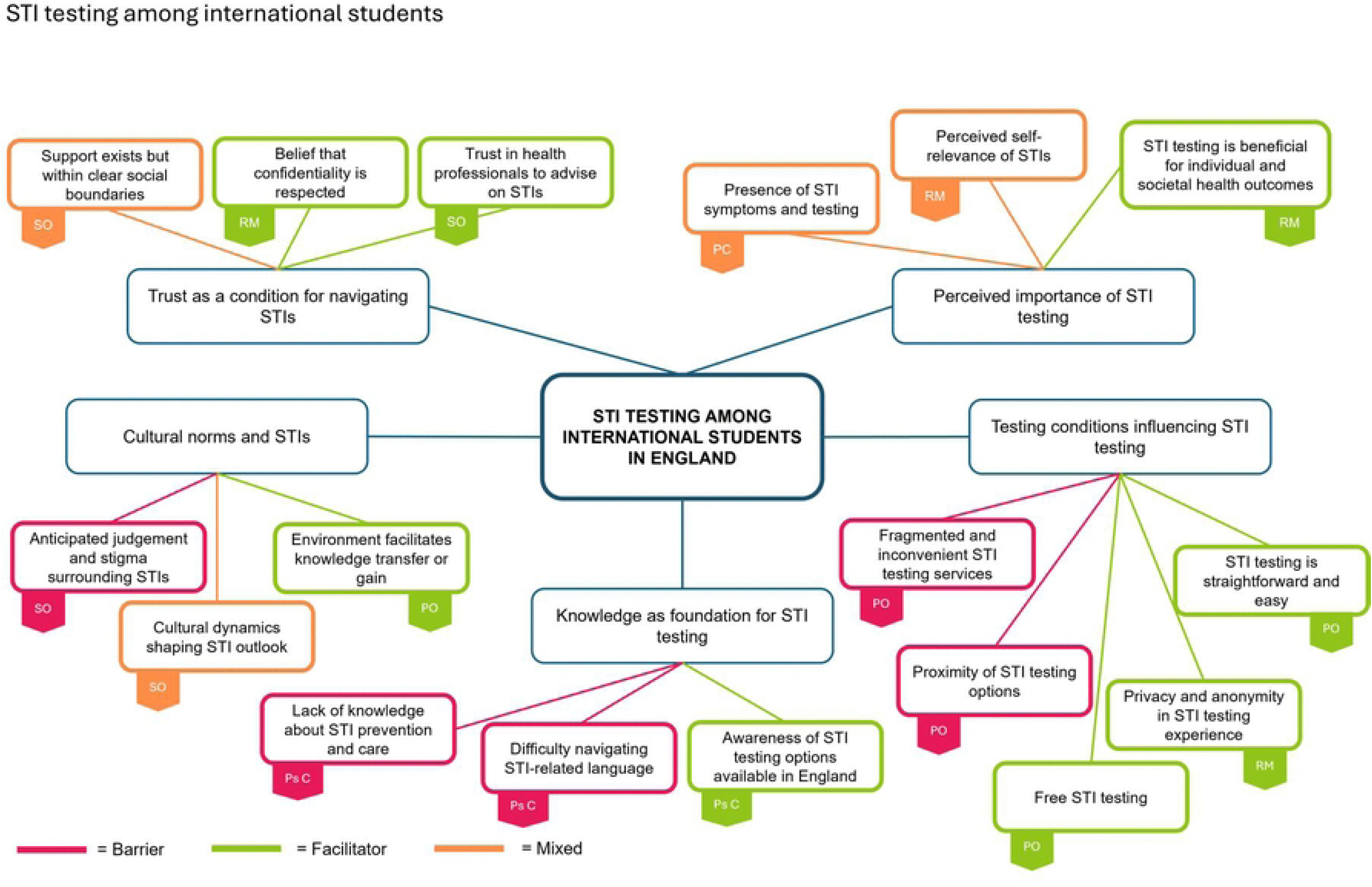
Overview of high and medium-priority themes. Overview of high and medium-priority themes clustered into 5 overarching narratives. Themes are colour-coded to indicate whether they acted as barriers (pink), facilitators (green), or were mixed – both barrier and facilitator (orange) on STI testing. Each theme is labelled with the corresponding COM-B domain. The abbreviations are as follows: Psychological Capability (Ps C), Physical Capability (PC), Reflective Motivation (RM), Social Opportunity (SO), and Physical Opportunity (PO).

## Trust as a condition for navigating STI testing

### Social Opportunity

### Trust in health professionals to advise on STIs

Most participants discussed trusting healthcare professionals to advise on STIs and sexual health, which also extended to friends and family. For some, the combination of professional expertise and close personal relationship fostered a sense of trust:

> “[…] [my friends] […] are professionals on the topic, yea so most of my friends are nurses because of my background […] we have experienced like several things together, so I know that I can talk about this with them like very easily because they understand” – P12 (25 to 34 years, male, born in the Americas, time lived in England between 6 months to 1 year)

Those with testing experience in England discussed how staff at SHSs helped create a comfortable and safe environment by treating them with confidentiality, further enabling trust.

#### Mixed

### Support exists but within clear social boundaries

Generally, participants viewed sexual health and STI topics as personal and private and not something that was openly spoken about. For instance, very few participants were aware of their friends’ STI testing behaviour:

> “My friends and I have never even talked about it […] maybe they’re doing it and they don’t feel confident about talking about it because of the […] perceived judgement they might receive. I don’t know, but it’s not very [much] talked about” – P7 (25 to 34 years, female, born in Nigeria, time lived in England between 6 months to 1 year)

While some participants noted an absence of STI support systems or chose not to engage with university services in favour of more trusted contacts, the majority felt they had sufficient support in place to facilitate testing. Participants were selective in their disclosure (preferring to disclose a negative test result rather than a positive one) and sharing sexual health information only with those deemed trustworthy and capable of offering unconditional support.

## Reflective Motivation

### Facilitators

#### Belief that confidentiality is respected

Some participants mentioned not having any concerns with confidentiality. Participants felt that organisations and healthcare staff respected their personal data and adhered to data protection regulations, which made them feel comfortable getting tested:

> “[…] I just feel that these kind of organisations and medical staff and stuff, they’re trained well [and] they probably have done their data protection training [and] all of these kinds of things, and they’re not go to just through some data on the street about my STI testing” – P8 (18 to 24 years, female, born in Europe, time lived in England between 3 to 5 years)

Some participants mentioned not having any concerns with confidentiality in medical procedures, in England or elsewhere.

## Cultural norms and STIs

### Social Opportunity

#### Barriers

##### Anticipated judgement and stigma surrounding STIs

Anticipated social stigma and negative impacts on relationships with friends and family were reasons for non-disclosure of testing. For other participants, it wasn’t so much about the act or the outcome of the test, but the implications associated with getting tested:

> “[…] I feel like [my parents] are telling me ‘you need to take good care of yourself’ which means if I got [an] STI I’m the one the be blamed […] because I’m not taking good care of myself […] they would ask me […] ‘why do you have unprotected sexual intercourse’ [or] ‘why are you not taking care of yourself’ […] I’m trying to avoid that judgement, so that’s why I didn’t tell them this time.” - P1(18 to 24 years, female, born in China, time lived in England between 3 and 5 years)

Most participants reported experiencing some degree of cultural taboo regarding sexual health, which impacted the way they internalised stigma and communicated about STIs:

> “[…] it might not be culturally accepted to go out in the open to discuss something like this […] society basically determines [how much people] are vocal about what they are going through […] I don’t think the stigma ever completely leaves, especially when the information is out there in the open. Back home [it may] be a difficult thing [to do].” – P3 (25 to 34 years, male, born in Nigeria, time lived in England 6 months to 1 year)

Three participants highlighted that while stigma still played a role, it did not prevent them from getting tested. Some were less concerned about stigma and instead described feeling uncomfortable or awkward; others were concerned about being recognised in clinics by peers.

#### Mixed

##### Cultural dynamics shaping STI outlook

Participants highlighted that England’s sexual health culture is more open than in their home countries’, with England-based social networks offering greater support for STI-related concerns. This openness was partly attributed to differences in sex education; Chinese participants specifically noted that sexual health lacks cultural acceptance in their home country, resulting in limited formal education on the topic, which reinforces discomfort around open discussion and leads individuals to seek STI knowledge privately:

> “[…] I think the stigma is very real […] I think this is rooted in wider culture of STIs, ‘cause in China, sex and anything related to sex [is] rarely spoken about, especially in public. So, traditional parents they offer no sex education at home, and for me, my parents never did. So, I had to […] figure things out myself. I [also] think the sex education material covered in school is also very minimal” – P5 (25 to 34 years, male, born in China, time lived in England between 1 and 2 years)

Participants’ perceptions about STI testing had evolved since living in England – they described being more open to getting tested and seeking support from peers on how to perform or access STI testing:

> “[…] I must say […] England is a lot more supportive about these kinds of things. So, people might [be more] likely to talk openly about these problems […] For now, [I] definitely feel more open than like, let’s say, six years ago… I can feel the gradual open phase within myself” – P2 (18 to 24 years, male, born in China, time lived in England between 3 and 5 years)

### Physical Opportunity

#### Facilitators

##### Environment facilitates knowledge transfer or gain

Participants mentioned learning about STIs and testing from a variety of sources while living in England. Some examples include bus stop advertisements, posters in sexual health clinics, NHS or official government websites, and social media (e.g., Instagram, Baidu, RedNote). Some participants observed that information is more widely available in England, than compared to their home country, particularly those from China and Nigeria:

> “I think there [is] more information available here, especially the information from NHS […] it’s much more better and available here […] there’s an official tool [to find sexual health clinics] […] in China, [if] I want to search for that, I can only find some things [are] not from government, it’s mostly private companies or from some non-profits” – P14 (25 to 34 years, male, born in China, time lived in England between 3 and 5 years)

## Knowledge as a foundation for STI testing

### Psychological Capability

#### Barriers

##### Lack of knowledge about STI prevention and care

Some participants discussed having limited knowledge about STIs in general, while others expressed not being able to interpret symptoms in the context of STIs:

> “[…] their names [are] one thing […] because I’m not familiar with that name, I’m not familiar with their symptoms as well. I’m not familiar with like the treatment and then the overall awareness of those disease[s]” – P1 (18 to 24 years, female, born in China, time lived in England between 3 and 5 years)

Participants also discussed not being aware of the different testing methods and other wider STI preventative interventions (e.g., vaccinations) available in England.

##### Difficulty navigating STI-related language

Participants expressed difficulties remembering, understanding and using technical STI terminology. STIs were viewed as having complex names which made them hard to remember; except HIV. Some participants felt they could not confidently communicate at a technical level with healthcare professionals:

> “[…] I think language is also a barrier because there are a lot of medical terms regarding the STI test and if I’m not very familiar with these terms, I cannot communicate with the doctors or staff working in the NHS.” - P5 (24 to 34 years, male, born in China, time lived in England between 1 and 2 years)

Other participants reported having challenges with knowing the English equivalent of STIs, some of them used translation apps during conversations with health professionals. This barrier was mentioned by most of the Chinese participants in our sample.

#### Facilitators

##### Awareness of STI testing options available in England

Students presented a good general awareness of the testing options available in England. Participants distinguished between different testing methods (i.e., blood test, urine or swab samples), and testing locations (e.g., sexual health clinics, GPs, and community-based organisations). Most knew that testing was free, including testing through online self-sampling kits. For some participants, a product of living in England was obtaining greater knowledge about STI testing through open conversations with peers allowing them to learn more about testing through informal social interactions. However, participants did not always explicitly link their knowledge to STI testing uptake:

> “[…] three months ago I have learned that I can have a free STI test in some […] sex clinic, so I just registered and took some tests […] I just learned from one of my friends […] this is the only resource, the first time I heard about this” – P13 (25 to 34 years, male, born in China, time lived in England 3 to 5 years)

## Testing conditions influencing STI testing

### Physical Opportunity

#### Barriers

##### Proximity of STI testing options

The cost (i.e., time and distance) associated with travelling to clinics was used as a reason why participants might not get tested:

> “[…] the time cost, it takes a lot of hours to get there, get back and also waiting there. It’s just time, it takes a lot […] [travelling to get tested in the future] will be a barrier for me” - P14 (25 to 34 years, male, born in China, time lived in England between 3 and 5 years)

##### Fragmented and inconvenient STI testing services

Participants who had previously used the English healthcare system found STI testing services complicated to navigate. Some were unable to secure appointments flexibly due to limited opening hours or because clinics were not accepting new patients. In other instances, participants delayed testing because they found the pre-appointment screening questions burdensome:

> “[when] I try to make an appointment, they ask detailed questions about my recent high-risk behaviour, but I didn’t want to pretend I had some […] so, I gave up” - P5 (25 to 34 years, male, born in China, time lived in England 1 to 2 years)

Two participants described feeling overwhelmed when accessing information online about testing. Online information was described as fragmented and confusing to understand:

> “[…] it just felt very overwhelming because I was confused, so I just left it […]it wasn’t very clear [what the conditions were] […]” - P7 (25 to 34 years, female, born in Nigeria, time lived in England between 6 months to a year)

#### Facilitators

##### Free STI testing

While most of the participants were aware that testing services were free in the UK, this did not often link to a decision to have a test, except for one participant:

> “[…] in the UK I think since this service is free, I think it sometimes encourages me to test” - P13 (25 to 34 years, male, born in China, time lived in England 3 to 5 years)

##### STI testing is straightforward and easy

Participants (either through experience or perception) viewed in clinic and at home testing as convenient and easy. Similar views were expressed by those who had previously used self-sampling test kits:

> “I’m far more likely to test when the test method is rapid, accurate, and easy to use” - P5 (25 to 34 years, male, born in China, time lived in England 1 to 2 years)

### Reflective Motivation

#### Facilitators

#### Privacy and anonymity in STI testing experience

Privacy and anonymity of testing was valued among participants because it helped reduce stigma:

> “I think the more private and convenient the process is, the more likely our international students are to use it […] if the university provide[d] some convenient and truly anonymous services for STI test[ing], I think this will definitely reduce the barriers for STI tests […] I think I prefer to choose the university service instead of NHS service [if this was the case]” – P4 (25 to 34 years, male, born in China, time lived in England 3 to 5 years)

For some participants this resolved concerns about being seen by other people. Discussion about the privacy of self-sampling tests was brought up predominantly by Chinese participants.

#### Perceived importance of STI testing

### Reflective Motivation

#### Facilitators

##### STI testing is beneficial for individual and societal health outcomes

Some viewed testing as important in maintaining individual mental and physical well-being, especially in terms of early detection and treatment of STIs:

> “[…] you only have one body […] you want to prevent getting serious illnesses […] it’s important to know whether you actually have an STI to get treatment the quickest you can to get rid of it […] you’re asking like is it important to breathe […] the benefits are for yourself.” - P8 (18 to 24 years, female, born in Europe, time lived in England 3 to 5 years)

Another benefit to testing was that it protected current and future sexual partners and was framed as a shared responsibility between partners. There was also mention of engaging in open conversation with partners about testing behaviour to help keep all parties safe:

> “[…] protect the people around you because you’re informed now of the situation. You’re not acting ignorantly anymore” - P7 (25 to 34 years, female, born in Nigeria, time lived in England 6 months to 1 year)

### Reflective Motivation

#### Mixed

##### Perceived self-relevance of STIs

Participants viewed themselves as being at low risk of STIs if they were sexually abstinent, in a monogamous relationship or frequently used condoms when engaging in sexual activity:

> “It [perception of risk] wouldn’t [change] cause I’m still going to make sure I’m protected. I’m aware of what is at stake, and I feel like I’m well informed when it comes to STIs and I am like consciously protecting myself so I wouldn’t put myself now or later in any position that puts me at risk.” – P7 (25 to 34 years, female, born in Nigeria, time lived in England between 6 months to 1 year)

In other cases, participants relied on trust and fidelity as indicators of low risk and safety:

> “[…] I have a very stable sex partner […] she’s my only sex partner […] [I] really believe in her, so, I don’t think I have any risk of STI” – P5 (24 to 35 years, male, born in China, time lived in England between 1 to 2 years)

Individual-level precautions and trust within interpersonal relationships contributed to creating a sense of safety, reducing the motivation to get tested for STIs. Conversely, participant’s perception of STI risk was influenced by changes in their sexual behaviour. For instance, participants expressed an intention to test more frequently if their sexual activity with new partners, with or without condoms, increased.

### Physical Capability

#### Presence of STI symptoms and testing

The presence of symptoms, or the lack thereof, acted as an indicator for testing behaviour. The influence of this factor varied for participants; some indicated that the presence of symptoms acted as a strong motivator for testing. However, in most cases, participants expressed an intention to test in a hypothetical future scenario where symptoms were present:

> “The thing is, if I am probably having symptoms or I see anything that looks like any symptoms, I’ll probably go to [the] GP and talk to [the] GP about it, and be referred, I guess, to the facility where I can do the testing.” - P3 (25 to 34 years, male, born in Nigeria, time lived in England between 6 months to 1 year)

### Behaviour Change Wheel Mapping

In Table 2, we mapped the themes that arose from barriers and mixed accounts to the Behaviour Change Wheel to inform a set of intervention functions (20) to facilitate STI testing among international students. Intervention functions were chosen based on best fit and in line with recommendations found in the literature.

**Table 2.**
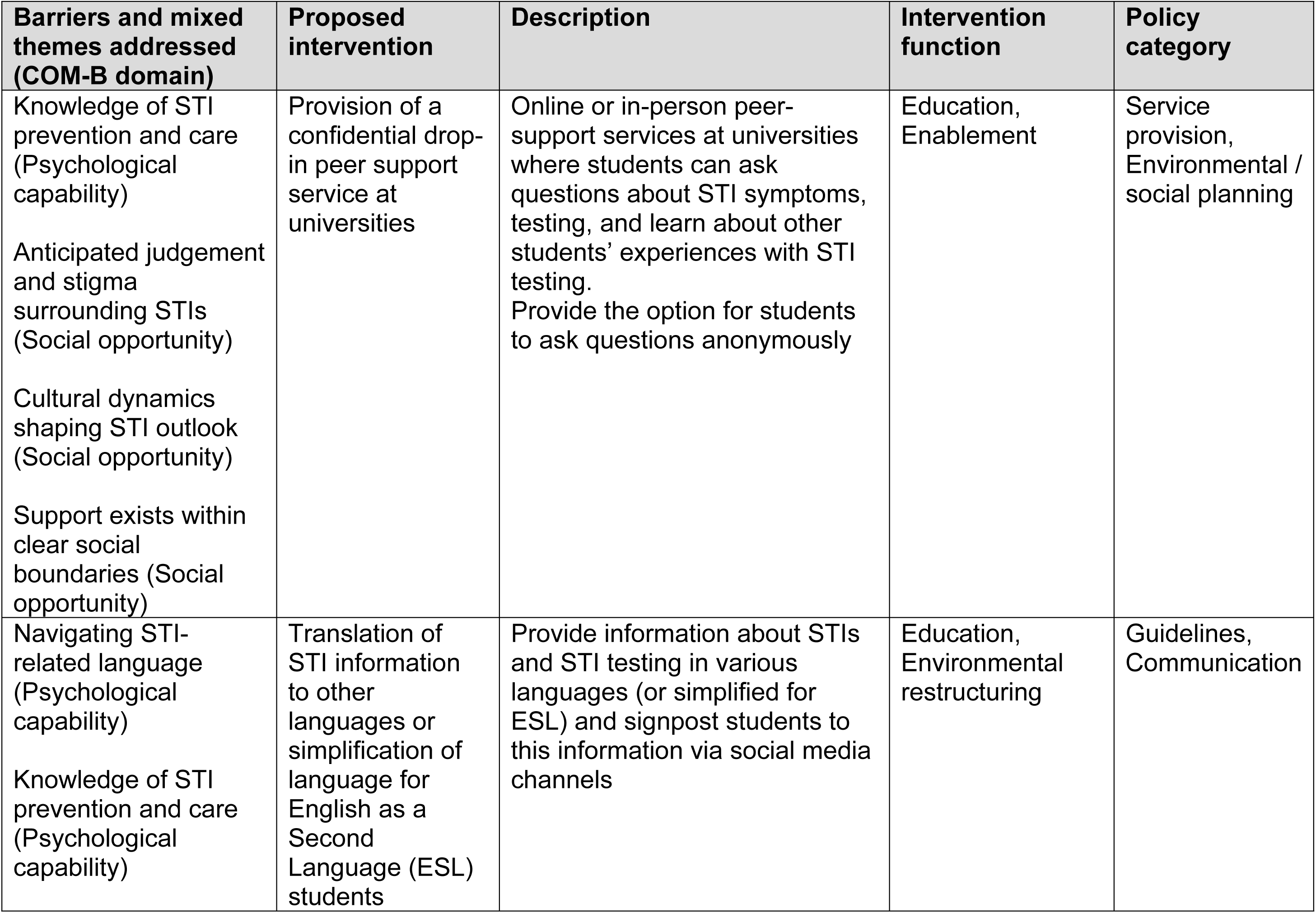
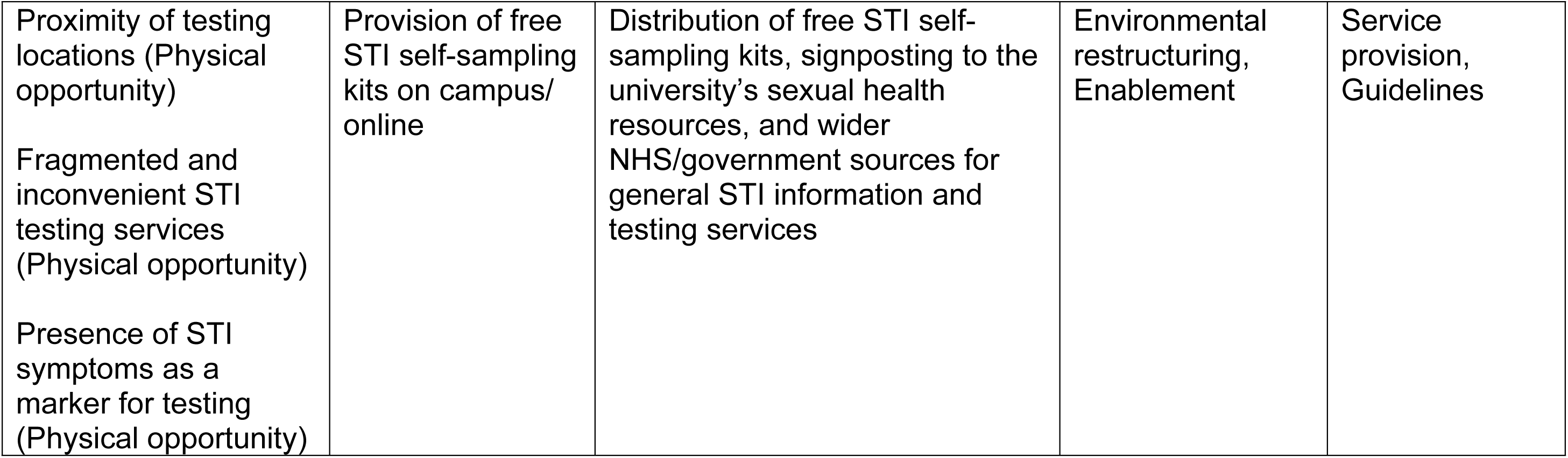
Example of key barriers and mixed themes mapped onto the BCW to inform interventions and policy.

| **Barriers and mixed themes addressed (COM-B domain)** | **Proposed intervention** | **Description** | **Intervention function** | **Policy category** |
|---|---|---|---|---|
| Knowledge of STI prevention and care (Psychological capability)<br>Anticipated judgement and stigma surrounding STIs (Social opportunity)<br>Cultural dynamics shaping STI outlook (Social opportunity)<br>Support exists within clear social boundaries (Social opportunity) | Provision of a confidential drop-in peer support service at universities | Online or in-person peer-support services at universities where students can ask questions about STI symptoms, testing, and learn about other students’ experiences with STI testing.<br>Provide the option for students to ask questions anonymously | Education, Enablement | Service provision, Environmental / social planning |
| Navigating STI-related language (Psychological capability)<br>Knowledge of STI prevention and care (Psychological capability) | Translation of STI information to other languages or simplification of language for English as a Second Language (ESL) students | Provide information about STIs and STI testing in various languages (or simplified for ESL) and signpost students to this information via social media channels | Education, Environmental restructuring | Guidelines, Communication |
| Proximity of testing locations (Physical opportunity)<br>Fragmented and inconvenient STI testing services (Physical opportunity)<br>Presence of STI symptoms as a marker for testing (Physical opportunity) | Provision of free STI self-sampling kits on campus/ online | Distribution of free STI self-sampling kits, signposting to the university’s sexual health resources, and wider NHS/government sources for general STI information and testing services | Environmental restructuring, Enablement | Service provision, Guidelines |

## Discussion

In this study, we applied the COM-B model (20) to identify the barriers and facilitators to STI testing among international students living in England. The confidentiality safeguards provided by SHSs, awareness of the availability of free STI testing at these services or online, and the perceived benefits of testing for individual and societal benefit were facilitators for STI testing. Conversely, technical STI-related language, limited knowledge of STIs, anticipated judgement and stigma linked to testing, fragmented service provision and poor proximity to services were barriers to STI testing in this group. The presence of STI symptoms, perceived self-relevance, different cultural dynamics and social support networks acted as both barriers and facilitators to STI testing. These findings highlight the need for clear, accessible language to describe STI-related symptoms and for clear public health messaging to reduce STI risk, especially among people for whom English is not their first language. These findings may apply to many young people, including British university students.

Participants expressed having limited knowledge about STIs, with some unable to interpret their own symptoms within an STI context. Others experienced challenges navigating England’s healthcare system, which contributed to postponed or delayed testing. Lack of knowledge about STIs and STI testing services is commonly reported in the literature as a barrier to engaging with STI testing (16, 22–24). In the present study, English not being one’s native language, constituted an additional barrier contributing to these knowledge gaps. Our participants noted that the names of STIs were hard to remember and found it hard to communicate at a technical level with medical professionals. This finding is consistent with evidence in the literature (14, 22, 25). This challenge might be explained by the fact that sexual health terms are not easily translated between languages, especially from Chinese to English (12, 25).

When compared to domestic students, several Australian studies have found international students to have lower levels of knowledge about sexual and reproductive health and STIs (12–14, 16), a disparity partly explained by differences in sex education. Discussion of sex and sexual health are considered taboo in many cultures (13, 15, 17), shaping the extent and quality of sex education young people receive (23, 26). This aligns with our participants’ perceptions, who felt domestic students were more open to discussing STIs, attributing this openness to a more sex-positive culture in England, supported by more comprehensive sex education and wider availability of sexual health information. In the absence of formal sex education, international students often turn to informal sources to fill these knowledge gaps such as social media, newspapers, and the internet (13, 23, 25–27). This is consistent with our findings: participants reported accessing general sexual health and STI testing information from social media platforms, and other trusted sources online (government, NHS, local councils, SHSs). Participants also stressed the importance of accessing reliable and targeted information from trusted sources, a finding also reported in the literature (12, 14, 15).

In our study, cultural norms in participants’ countries of origin did not appear to discourage participants from getting tested; rather, a cultural tension emerged more specifically around disclosing testing behaviours to others. Although some participants felt that living in England broadened their mindset through exposure to a more open, sex-positive environment with more supportive systems in place, this openness did not necessarily extend to how comfortable participants felt sharing their testing status with friends and family from their country of origin. This tension became apparent when disclosing a positive result: while participants generally had no issue disclosing a negative test, they were far more cautious about disclosing a positive one, given its association with unsafe sex and having multiple partners. Similar concerns regarding testing behaviour were observed in young people living in the UK, across Europe, and in the United States (6, 10, 28–30).

Participants managed this tension through selective disclosure and information-seeking with friends and family, choosing to share their testing status with selected friends and family members, and trusted health professionals, while withholding this information from those they feared might judge their behaviour. Social norms, and the associated stigma and judgement from others, appear to be a key driver of this behaviour, a pattern reported in the literature (6, 15, 17, 26, 28). In our findings testing attitudes were largely unaffected, which may reflect the predominance of students doing health-related degrees in our sample. This contrasts with previous research demonstrating that STI-related shame and stigma are associated with more negative attitudes towards testing and can act as barriers to testing uptake in other populations (10, 30). This suggests that host-country norms can shift attitudes toward the act of testing while home-country stigma continues to shape testing disclosure.

Similar experiences of shame and stigma have been reported in the wider literature on young people in general. Barriers to testing reflected local social environment, institutions, shame, peer influence, and gendered expectations rather than home-culture carryover. A number of these barriers centred on fears of others’ reactions: fear of parents’ or partners’ reactions and feeling judged by others more broadly (11), as well as anticipated judgement from peers specifically (8). Others reflect self-directed shame and negative self-evaluation, informed by part experiences and anticipated feelings about testing (6), including fears of receiving negative feedback about one’s sexual behaviour or being viewed as “unclean” (8).

### Practical implications

Based on the behavioural diagnosis and the Behaviour Change Wheel mapping (Table 2) we suggest three interventions to address the key barriers and mixed accounts described by participants. Culturally driven stigma is a deeper-rooted structural barrier that is complex to address; however, offering a peer support service at universities could, in part, help address the social opportunity and psychological capability barriers discussed previously. A peer-support service could be available for students to ask peers outside of their immediate social networks about accessing and navigating STI testing services and learn of others’ experiences with STI testing. Offering an online peer support service, where students are also able to ask their questions anonymously, may help address privacy concerns and selective disclosure approaches used to maintain confidentiality. There are several existing community programmes offered by London-based organizations delivering services such as counselling and peer support (31), that could be used as models. Delivering education via peer facilitators that helps people understand where and how to get tested are considerations for optimisation outlined by the STI Prioritisation Framework for England (32).

Translating STI information into multiple languages can remove the burden of interpreting unfamiliar medical terminology, limit the risk of misunderstanding symptoms, or testing procedures. While international students attending university in England must meet a specific English Language Proficiency standard, this is unlikely to account for medical or health-specific jargon. Enabling international students to access information in their native language can help remove a language barrier while helping close pre-existing gaps in sexual health knowledge. This recommendation aligns with those found in the literature (13, 33, 34) and could be modelled after the multilingual toolkit produced by the New South Wales Multicultural HIV and Hepatitis Service (35). Public-facing materials to promote vaccination programmes in England, including those targeted to university students, are provided in multiple languages (36).

Many sexual health services in England currently offer at-home STI self-sampling kits (37), which is a popular option (38, 39). However, participants continued to experience access barriers which could be addressed at universities by providing STI self-sampling kits to students in need of STI testing. The provision of these kits can address proximity and fragmentation barriers, as it would remove the need to locate, travel, or navigate multiple services. Furthermore, the information shared in these kits can improve understanding of both symptomatic and asymptomatic STIs, helping people recognise the potential relevance of STI testing to their own circumstances. Fresher’s week, an orientation period for new university students featuring social events (40), could be an opportune time to raise awareness of sexual health provision and signpost to National Health Service and university healthcare information and resources. Early insights from this study were used to produce STI public health messaging targeted at university students in the England by UKHSA.

## Strengths and Limitations

A key strength of this study is the qualitative study design that allowed for collecting direct and in-depth accounts from international students living in England, a subgroup of younger people with distinct health needs, on the barriers and facilitators to STI testing. Our study reached a diverse sample of international students, including those from countries with high gonococcal ceftriaxone resistance. Additionally, the application of the COM-B model enabled a theory-informed and systematic assessment of behavioural determinants and their mapping to intervention and policy approaches through the Behaviour Change Wheel. A potential limitation is that participants in our sample predominantly came from a health background. Given this, participants might have a greater awareness of sexual health topics and be more health conscious. They might have also felt inclined to express attitudes they perceived as professionally or socially expected, introducing the possibility of social desirability bias. The predominance of postgraduate students in our sample may limit the transferability of the findings to younger and undergraduate student populations, whose life circumstances, perceived susceptibility to STIs, and STI-testing behaviours may differ. Conversely, the study may not have adequately captured students who are less engaged with sexual health services, whose attitudes and experiences regarding STI testing may differ from those represented in the sample. Furthermore, this study largely captured views of participants who had previous testing experience, either in England or in their respective home countries. This may make them more comfortable discussing testing behaviour, more likely to express positive intentions to test, and less likely to report some anticipated barriers that first-time testers might feel more acutely. Lastly, we recruited only participants from a university in London, and the barriers and facilitators to STI testing may vary in other cities of England.

## Conclusion

This study contributes to the limited evidence base on barriers and facilitators to STI testing in international students in England, a large and heterogeneous group of people of different cultural backgrounds, with many at the age of forming new or casual sexual partnerships. Insights from this research have been used by UKHSA to optimise STI public health messaging for all university students in England (41), and the accompanying social media assets used for STI health promotion. The evidence from our study can be used to create culturally appropriate resources to raise awareness of STI-related symptoms and, among those with a need, promote STI testing.

## Data Availability

Data supporting this study cannot be made available due to participant consent limitations.

## Acknowledgements

We want to thank the following people for their contributions to this study: All participants who generously shared their time and insights, our PPIE advisors for their input into the cultural appropriateness and accessibility of the study materials, and to the university staff who kindly supported the distribution of recruitment materials to students.

